# Characteristics and risk factors for SARS-CoV-2 among children in Italy: a cross-sectional study in 20 pediatric centers

**DOI:** 10.1101/2021.03.17.21253610

**Authors:** Marzia Lazzerini, Idanna Sforzi, Sandra Trapani, Paolo Biban, Davide Silvagni, Ilaria Mariani, Giovanna Villa, Jessica Tibaldi, Luca Bertacca, Enrico Felici, Giuseppina Perricone, Roberta Parrino, Claudia Gioè, Sara Lega, Mariasole Conte, Federico Marchetti, Annamaria Magista, Paola Berlese, Stefano Martelossi, Francesca Vaienti, Enrico Valletta, Margherita Mauro, Roberto Dall’Amico, Silvia Fasoli, Antonio Gatto, Antonio Chiaretti, Danica Dragovic, Paola Pascolo, Chiara Pilotto, Ilaria Liguoro, Elisabetta Miorin, Francesca Saretta, Gianluca Trobia, Antonella Di Stefano, Azzurra Orlandi, Fabio Cardinale, Riccardo Lubrano, Alessia Testa, Marco Binotti, Valentina Moressa, Egidio Barbi, Benedetta Armocida on behalf of COVID-19 Italian Pediatric Study Network

## Abstract

**Background:** No study has described factors associated with COVID-19 diagnosis in children.

**Aim:** Describe characteristics and risk factors for COVID-19 diagnosis in children tested in 20 pediatric centers across Italy.

**Methods:** Cases aged 0-18 years tested for SARS-CoV-2 between February 23 and May 24 2020 were included. Our primary analysis focused on children tested because of COVID-19 suggestive symptoms.

**Results:** Among 2494 children tested for SARS-CoV-2, 2148 (86.1%) had symptoms suggestive of COVID-19. Clinical presentation of SARS-CoV-2 included - beside fever (82.4%) and respiratory signs or symptoms (60.4%) – also gastrointestinal (18.2%), neurological (18.9%), cutaneous (3.8%) and other flu-like presentations (17.8%). In multivariate analysis, factors significantly associated with SARS-CoV-2 were: exposure history (adjusted OR 39.83 95%CI 17.52-90.55 p<0.0001), cardiac disease (adjusted OR 3.10 95%CI 1.19-5.02 p<0.0001), fever (adjusted OR 3.05 % 95% CI 1.67-5.58 p=0.0003), and anosmia/ageusia (OR 4.08 95%CI 1.69 −9.84 p=0.002). Among 190 (7.6%) children diagnosed with SARS-CoV-2, only four (2.1%) required respiratory support and two (1.1%) were admitted in ICU, while 100% recovered.

**Conclusion:** Recommendations for SARS-CoV-2 testing in children should be updated based on the evidence of broader clinical features. Exposure history, fever, and anosmia/ageusia are strong risk factors for COVID-19 in children, while other symptoms don’t seem helping discriminating in between the SARS-CoV-2 positive and the negative cases. This study confirm that COVID-19 is a mild disease in the general population of children in Italy. Further studies are needed to understand the risk, clinical spectrum and outcomes of COVID-19 in children with specific preexisting conditions.

## INTRODUCTION

The worldwide outbreak of severe acute respiratory syndrome coronavirus 2 (SARS-CoV-2), which emerged in Wuhan China in December 2019, rapidly affected Italy [1]. The first confirmed cases of coronavirus disease (COVID-19) in Italy were diagnosed on January 29, 2020. The Italian government declared a “state of emergency” by 31^st^ January 2020 [2], and by 24^th^ May 2020 a total of 229.858 cases of COVID-19 had been diagnosed across the country [3].

From the very beginning of the pandemic, data suggested that children are less affected than adults by COVID-19 [4-10]. However, timely diagnosis of COVID-19 is not only important for the single individual; it is crucial to prevent the spread of the pandemic. A better understanding of the predictors of positive SARS-CoV-2 test results may facilitate timely case finding and contact tracing and thus largely contribute to control the pandemic. It may also improve organization of care in settings were diagnostic facilities are available but still require a considerable processing time, where diagnostic facilities are lacking, and where diagnosis, in absence of other tools, may need to be based on clinical features alone.

Several systematic reviews have so far synthetized the clinical features and outcomes of pediatric cases with a confirmed infection by SARS-CoV-2 [11-17], but none published study yet explored the risk factors associated with a positive diagnosis of “COVID 19” among children undergoing a diagnostic test. It is currently unknown whether risk factors explored in adults [18-21] - such as exposure history, obesity, lymphocytopenia, or ground glass opacity at lung X-ray [19] - apply for children. So far, only one unpublished report analyzed variables associated with increased odds of SARS-CoV-2 diagnosis in the pediatric population, but the sample was very small (77 cases) and the cases, retrospectively enrolled among hospitalized children, were not representative of the general pediatric population tested for SARS-CoV-2 [22].

This study aimed at describing the characteristics of pediatric patients tested for SARS-CoV-2 during the early phase of the pandemic in 20 centers across Italy, and at exploring risk factors associated with positive diagnosis of COVID-19. The sample was divided in the followings subgroups of children: a) children tested because of symptoms suggestive of COVID-19; b) asymptomatic children tested because of contact with a COVID-19 positive case; c) hospitalized children tested because of hospital screening programs.

## METHODS

### Study designs and participants

This cross-sectional study is reported according to STROBE guidelines [23]. Data were collected through a collaborative research network coordinated by the World Health Organization (WHO) Collaborating Center in Maternal and Child health of the Institute for Maternal and Child Health IRCCS Burlo Garofolo, Trieste, Italy. Cases aged 0-18 years tested for SARS-CoV-2 in the period between ^February^ 23 and May 24, 2020 in any of the 20 pediatric centers participating in the network were included in the study.

National recommendations on SARS-CoV-2 testing did not change during the study period, and indicated testing for: 1) contacts of COVID-19 positive cases; 2) cases of severe acute respiratory distress syndrome; 3) cases with either fever, cough or difficulty breathing and absence of another etiology that could fully explain the clinical presentation [24]. In addition, based on the local epidemiology and on emerging evidence on COVID-19 [25-27], some facilities implemented local policies of testing all children hospitalized and/or children with gastrointestinal or cutaneous symptoms, such as vasculitis, during the study period.

SARS-CoV-2 infection was diagnosed, in line with national recommendations, using nasal or nasopharyngeal swab specimens collected by trained personnel, and tested for SARS-CoV-2 nucleic acid in regional referral laboratories using WHO-recommended real-time reverse-transcriptase polymerase-chain-reaction (*RT-PCR*) assays.

### Data collection and management

Data were collected with a standardized, field-tested on-line anonymous form, previously utilized for another study [12], and further optimized and adapted for the purpose of this study. The form collected variables to classify children in the following pre-defined categories: a) children tested because of symptoms suggestive of COVID-19; b) asymptomatic children tested because of contact with a COVID-19 positive case; c) hospitalized children tested because of hospital screening programs. It included information on socio-demographic and clinical characteristics, diagnostic examinations, type of treatments, and outcomes. Both closed and open questions were utilized. Data were obtained from official medical records and entered in the form by clinical staff in charge of case management at each facility level. Information for health workers on how to complete the form was embedded in the form itself. Data collection forms where checked in real time for internal consistency or missing data by trained personnel. Additional cross-check and data cleaning were conducted before data analysis by two expert biostatisticians (IM, BA).

### Study variables

We included in this study socio-demographic and clinical characteristics, diagnostic examinations, type of treatments, and outcomes variables. For children tested because of symptoms suggestive of COVID-19, disease severity was classified using a pre-defined objective criterion adapted from previously published classification [12], as reported in **Supplementary Table 1**. Tachypnea and tachycardia where defined as detailed in **Supplementary Table 2**. The outcome variable for multivariate analysis was testing positive for SARS-CoV-2.

### Statistical analysis

Categorical variables were reported as absolute numbers and percentages. Continuous variables were expressed as means and standard deviations (SD) or as median and inter-quartile ranges (IQR), if not normally distributed. We tested for associations between individual covariates and the outcome of a positive SARS-CoV-2 swab using χ^2^ test or Fisher’s exact test, as appropriate. Variables with a significant univariate relationship with the outcome, available in the whole sample, unless collinear, were included in a generalized estimating equations (GEE) logistic regression model using a compound symmetry covariance structure within centers. The GEE model accounts for correlation between patients who refer to the same center. We performed separate analyses in the three subgroups: a) children tested because of symptoms suggestive of COVID-19; b) asymptomatic children tested because of a contact with a COVID-19 positive case; c) hospitalized children tested because of hospital screening programs. The analyses on children tested because of symptoms suggestive of COVID-19 were predefined as our primary analyses, while the analyses in the other two subgroups were considered secondary analyses. An exploratory subgroup analysis was performed on disease severity by age group and sex in patients with symptoms suggestive of COVID-19. We also performed secondary analyses to describe variation across centers in the rate of children with positive SARS-CoV-2-testing. The significance level was set at 0.05 (two-tailed test). Data were analyzed with STATA 14 and SAS 9.4.

## Ethical considerations

The study was approved by the Institutional Review Board of the Institute for Maternal and Child Health IRCCS Burlo Garofolo, Trieste, Italy (reference number 01/2020 25.03.2020). Data were collected in an anonymous way, analyzed and reported only in aggregate form. Given the purely descriptive and retrospective nature of the study, informed consent was waived.

## RESULTS

During the study period, 2494 children were tested for SARS-CoV-2 across 20 centers **(Figure 1)**. Geographical case distribution is depicted in **Supplementary Figure 1**. Out of the total sample, 2148 (86.1%) children were tested because of symptoms suggestive of COVID-19; 52 (2.1%) asymptomatic children were tested because of contact with a COVID-19 positive case; 294 (11.8%) children were tested within hospital screening programs. Among all the tested children, 190 (7.6%) resulted positive. The percentage of positive cases was significantly higher in those tested because of a COVID-19 positive contact (rate of COVID-19 positive 51.9%) than in those tested because of symptoms (rate of COVID-19 positive 7.4%, p<0.0001) or in hospital screening programs (rate of COVID-19 positive 2.1%, p<0.0001) **(Figure 1)**.

**Figure 1.** Study flow diagram.

The clinical presentations of SARS-CoV-2 positive cases tested because of symptoms included – besides fever and/or respiratory signs or symptoms - gastrointestinal, neurological, and dermatological manifestations and other flu-like features (Figure 2). Specifically, 131 children (82.4%) had fever, which presented as the only symptom in 26 (25.2%) and 96 (60.4%) had respiratory signs/symptoms, which presented alone in nine (5.7%). Neurological symptoms - such as convulsion, irritability, headache, anosmia/ageusia-were observed in 30 (18.9%), in one child as the only symptom. Non-specific flu-like symptoms, such as muscular-articular pains, nausea, poor appetite, were reported in 27 (17.0%), always in combination with other clinical signs. Six children were tested due to cutaneous signs such as vasculitis and pseudo-chilblains on fingertips and toes, always in association with other symptoms of any type (either fever, respiratory, neurological, flu-like or gastrointestinal).

**Figure 2.** Clinical presentation of SARS-CoV-2 positive children. Abbreviations: GI = gastro-intestinal; Resp = respiratory; Neuro = Neurological.

When comparing children based on the results of swab testing (**Table 1**), children SARS-CoV-2 positive were more often in the stratum of 10 to 18 years compared to the SARS-CoV-2 negative (54.1% vs 26.0%, p<0.001). No difference by sex was observed. A history of COVID-19 positive contact was strongly associated with an increased risk of positive swab (79.2% vs 6.1%, p<0.001). Similarly, having a relative with respiratory symptoms was strongly associated with SARS-CoV-2 positive test results (72.3% vs 11.5% p<0.001). Both positive and negative children had a non-negligible rate of comorbidities (17.6% vs 16.4%, p=0.7), with cardiac diseases being slightly more frequent in the group testing positive for SARS-CoV-2 (5.7% vs 2.1% p=0.005).

**Table 1.** Socio-demographic characteristics of children tested because of symptoms suggestive for COVID-19.

|  | Positive swab<br>N=159 |  | Negative swab<br>N=1989 |  |  |
| --- | --- | --- | --- | --- | --- |
|  | n | % | n | % | p-values |
| **Age groups** |  |  |  |  |  |
| < 6 months | 19 | 12.0 | 159 | 8.0 | 0.082 |
| 6-24 months | 17 | 10.7 | 472 | 23.7 | **<0.001** |
| 2-9 years | 37 | 23.3 | 836 | 42.0 | **<0.001** |
| 10-18 years | 86 | 54.1 | 517 | 26.0 | **<0.001** |
| Missing | 0 | 0 | 5 | 0.3 | 1.000 |
| **Sex** |  |  |  |  |  |
| Male | 77 | 48.4 | 1108 | 55.7 | 0.076 |
| Female | 82 | 51.6 | 880 | 44.2 | 0.074 |
| Missing | 0 | 0 | 1 | 0.1 | 1.000 |
| **Contact with COVID-19 positive** | 126 | 79.2 | 122 | 6.1 | **<0.001** |
| **Relatives with respiratory symptoms** | 115 | 72.3 | 229 | 11.5 | **<0.001** |
| **Any co-morbidity** | 28 | 17.6 | 327 | 16.4 | 0.702 |
| **Type of comorbidities** |  |  |  |  |  |
| Malformation, disabilities, neuromuscular diseases | 5 | 3.1 | 81 | 4.1 | 0.566 |
| Cardiac diseases | 9 | 5.7 | 42 | 2.1 | **0.005** |
| Asthma | 6 | 3.8 | 60 | 3.0 | 0.593 |
| Others Respiratory diseases/conditions | 0 | 0 | 17 | 0.9 | 0.631 |
| Primary immunodeficiencies | 1 | 0.6 | 12 | 0.6 | 1.000 |
| Secondary immunodeficiencies | 1 | 0.6 | 41 | 2.1 | 0.365 |
| Obesity | 1 | 0.6 | 12 | 0.6 | 1.000 |
| Diabetes | 1 | 0.6 | 2 | 0.1 | 0.206 |
| Psychiatric disorders | 1 | 0.6 | 21 | 1.1 | 1.000 |
| Other | 9 | 5.7 | 97 | 4.9 | 0.659 |

When compared for disease severity at presentation (**Table 2**), there were no significant differences between children who were SARS-CoV-2 positive and those negative, with most cases having a mild presentation (78% and 70.8%, respectively, p=0.053) and an equal number of cases having a severe (3.1% vs 6.7%, p=0.091) or critical presentation (1.3% vs 1.0%, p=0.665) in each group.

**Table 2.** Clinical presentation and outcomes of children tested because of symptoms suggestive for COVID-19.

|  | Positive swab<br>N=159 |  | Negative swab<br>N=1989 |  |  |
| --- | --- | --- | --- | --- | --- |
|  | n | % | n | % | p-values |
| **Disease severity at presentation** |  |  |  |  |  |
| Asymptomatic | 8 | 5.0 | 125 | 6.3 | 0.528 |
| Mild | 124 | 78.0 | 1408 | 70.8 | 0.053 |
| Moderate | 20 | 12.6 | 304 | 15.3 | 0.359 |
| Severe | 5 | 3.1 | 133 | 6.7 | 0.091 |
| Critical | 2 | 1.3 | 19 | 1.0 | 0.665 |
| **Symptoms and signs at presentation** |  |  |  |  |  |
| Fever | 131 | 82.4 | 1355 | 68.1 | **<0.001** |
| Respiratory symptoms, any | 96 | 60.4 | 1325 | 66.6 | 0.110 |
| Respiratory distress | 12 | 7.5 | 255 | 12.8 | 0.052 |
| Rhinorrhea | 32 | 20.1 | 372 | 18.7 | 0.659 |
| Dry cough | 51 | 32.1 | 452 | 22.7 | **0.007** |
| Productive cough | 7 | 4.4 | 185 | 9.3 | **0.037** |
| Sore throat | 36 | 22.6 | 881 | 44.3 | **<0.001** |
| Strep throat | 2 | 1.3 | 106 | 5.3 | **0.024** |
| Conjunctivitis | 8 | 5.0 | 60 | 3.0 | 0.163 |
| Apnea | 0 | 0 | 4 | 0.2 | 1.000 |
| Thoracic pain | 6 | 3.8 | 44 | 2.2 | 0.209 |
| **Gastrointestinal symptoms, any** | 29 | 18.2 | 574 | 28.9 | **0.004** |
| Vomiting | 16 | 10.1 | 365 | 18.3 | **0.009** |
| Diarrhea | 18 | 11.3 | 293 | 14.7 | 0.240 |
| **Neurological symptoms, any** | 30 | 18.9 | 175 | 8.8 | **<0.001** |
| Asthenia | 10 | 6.3 | 38 | 1.9 | **<0.001** |
| Headache | 13 | 8.2 | 79 | 4.0 | **0.012** |
| Anosmia/ageusia | 13 | 8.2 | 10 | 0.5 | **<0.001** |
| Convulsion | 2 | 1.3 | 49 | 2.5 | 0.583 |
| Hyperactivity | 1 | 0.6 | 12 | 0.6 | 1.000 |
| **Cutaneous presentations, any** | 6 | 3.8 | 159 | 8.0 | 0.054 |
| Skin manifestations | 6 | 3.8 | 158 | 7.9 | 0.057 |
| Vasculitis | 0 | 0 | 11 | 0.6 | 1.000 |
| **Unspecific flu-like presentations, any** | 27 | 17.0 | 303 | 15.2 | 0.557 |
| Muscle or joint pains | 18 | 11.3 | 71 | 3.6 | **<0.001** |
| Nausea | 0 | 0 | 13 | 0.7 | 0.616 |
| Inappetence | 15 | 9.4 | 237 | 11.9 | 0.349 |
| Lymphadenitis | 8 | 5.0 | 158 | 7.9 | 0.186 |
| **Other symptoms, any** | 23 | 14.5 | 528 | 26.6 | **0.001** |
| Abdominal pains | 11 | 6.9 | 269 | 13.5 | **0.017** |
| Oral manifestations<br>(gingivostomatitis, aphthae) | 2 | 1.3 | 54 | 2.7 | 0.433 |
| Dental problems | 1 | 0.6 | 6 | 0.3 | 0.417 |
| Urogenital disorders | 0 | 0 | 10 | 0.5 | 1.000 |
| Ear problems | 0 | 0 | 32 | 1.6 | 0.166 |
| Others | 3 | 1.9 | 41 | 2.1 | 1.000 |
| **Vital parameters at presentation** |  |  |  |  |  |
| Tachycardia | 12/61 | 19.7 | 294/1489 | 19.7 | 0.989 |
| Tachypnea | 4/34 | 11.8 | 187/827 | 22.6 | 0.204 |
| **Oxygen saturation level at presentation** |  |  |  |  |  |
| 91-92 | 2/66 | 3.0 | 15/1575 | 0.1 | 0.147 |
| ≤90 | 1/66 | 1.5 | 21/1575 | 1.3 | 0.597 |
| **Clinical examination at presentation** |  |  |  |  |  |
| **Lungs auscultations** |  |  |  |  |  |
| Negative | 69/86 | 80.2 | 1486/1826 | 81.3 | 0.810 |
| Crackles | 4/86 | 4.7 | 186/1826 | 10.2 | 0.099 |
| Wheezing | 3/86 | 3.5 | 120/1826 | 5.6 | 0.366 |
| Absent breath sounds | 4/86 | 4.7 | 111/1826 | 6.1 | 0.816 |
| **Laboratory test<sup>1</sup>** |  |  |  |  |  |
| White blood cell count <5.5 (×10<sup>9</sup>/L) | 17/50 | 34.0 | 109/809 | 13.5 | **<0.001** |
| Lymphocyte count <1.2 (×10<sup>9</sup>/L) | 8/41 | 19.5 | 75/559 | 13.4 | 0.275 |
| Neutrophil <1.50 (×10<sup>9</sup>/L) | 6/47 | 12.8 | 50/743 | 6.7 | 0.118 |
| C-reactive protein > 1 gr/dl | 29/50 | 58.0 | 589/760 | 77.5 | **0.002** |
| Erythrocyte sedimentation rate > 20 mm/h | 2/4 | 50.0 | 34/64 | 53.3 | 1.000 |
| Aspartate aminotransferase > 50 (U/L) | 7/35 | 20.0 | 56/434 | 12.9 | 0.234 |
| Alanine aminotransferase > 45 (U/L) | 4/46 | 8.7 | 74/692 | 10.7 | 0.808 |
| D dimer >0.5 (µg/mL) | 2/4 | 50.0 | 24/46 | 52.2 | 1.000 |
| **Chest X-ray** | 27 | 17.0 | 313 | 15.7 | 0.679 |
| Negative | 8/27 | 29.6 | 106/313 | 33.9 | 0.655 |
| Ground glass opacities | 7/27 | 25.9 | 71/313 | 22.7 | 0.701 |
| Focal consolidation | 3/27 | 11.1 | 77/313 | 24.6 | 0.155 |
| Other description | 9/27 | 33.3 | 59/313 | 18.8 | 0.071 |
| **Lung ultrasound** | 5 | 3.1 | 58 | 2.9 | 0.806 |
| Negative | 1/5 | 20.0 | 18/58 | 38.0 | 1.000 |
| B-lines in various pattern | 2/5 | 40.0 | 30/58 | 51.7 | 0.672 |
| Focal consolidation | 0 | 0 | 5/58 | 8.6 | 1.000 |
| Other description | 1/5 | 20.0 | 1/58 | 1.7 | 0.154 |
| **Lung CT scan** | 5 | 3.1 | 13 | 0.7 | **0.008** |
| Negative | 0/5 | 0 | 2/13 | 15.4 | 1.000 |
| Ground glass opacities | 3/5 | 60.0 | 6/13 | 46.2 | 1.000 |
| Focal consolidation | 1/5 | 20.0 | 3/13 | 23.1 | 1.000 |
| Other description | 0/5 | 0 | 2/13 | 15.4 | 1.000 |
| **Hospitalised** | 45 | 28.3 | 602 | 30.3 | 0.603 |
| **Respiratory support<sup>1</sup>** | 4 | 2.5 | 73 | 3.7 | 0.656 |
| Oxygen | 3/45 | 6.7 | 54/602 | 9.0 | 0.788 |
| High flow oxygen | 2/45 | 4.4 | 19/602 | 3.2 | 0.651 |
| Noninvasive ventilation | 1/45 | 2.2 | 4/602 | 0.7 | 0.303 |
| Mechanical ventilation | 0/45 | 0 | 11/602 | 1.8 | 1.000 |
| **Cases in ICU** | 2 | 1.3 | 11 | 0.6 | 0.250 |
| **Outcome** |  |  |  |  |  |
| Cured | 159 | 100 | 1981 | 99.6 | 0.500 |
| Referred | 0 | 0 | 7 | 0.4 | 0.460 |
| Dead | 0 | 0 | 1 | 0.1 | 1.000 |

Abbreviations: ICU= intensive Care Unit
1 Note: available in a subsample of cases

Fever was highly prevalent in both groups, significantly more in SARS-CoV-2 positive children (82.4% vs 68.1%, p< 0.001). Respiratory symptoms were highly prevalent in both groups (60.4% vs 66.6%, p=0.110) with some differences: dry cough was more frequent in the group of SARS-CoV-2 positives (32.1% vs 22.7%, p=0.007), whereas sore throat and strep throat were less frequent in this group compared to the negatives (22.6% vs 44.3%, p<0.001 and 1.3% vs 5.3%, p=0.024 respectively). Respiratory distress was less frequent in the group testing positive for SARS-CoV-2 than in the group testing negative, although the difference was not statistically significant (7.5% vs 12.8%, p=0.052).

Gastrointestinal symptoms (18.2% vs 28.9%, p=0.004), and other symptoms (14.5% vs 26.6%, p=0.001) were significantly less frequent in the group of SARS-CoV-2 positives, while the opposite was true of neurological symptoms (18.9% vs 8.8%, p<0.001) and muscle or joint pains (11.3% vs 3.6%, p<0.001).

Vital parameters, as well as oxygen saturation levels and lung auscultation were not significantly different between the two groups. Lymphocytopenia was significantly more frequent in the SARS-CoV-2 positive group (34.0% vs 13.5%, p<0.001), while elevated C-reactive protein was more frequent in the SARS-CoV-2 negative group (58.0% vs 77.5%, p=0.002). Findings at chest-X-ray, lung ultrasound and lung CT scan were not significantly different among the two groups, with equal prevalence of ground glass opacities (respectively 25.9% vs 22.7%, p=0.7, 40.0% vs 51.7% p=0.67).

The frequencies of hospitalized cases (28.3% vs 30.3%, p=0.60) and those admitted to intensive care (ICU) (1.3% vs 0.6%, p=0.25) were not significantly different between the SARS-CoV-2 positive and negative children. Need and type of respiratory support was also not significantly different. Final outcomes did not differ between groups, although in the SARS-CoV-2 negative group one death was observed.

### Multivariate analysis

In multivariate analysis, factors significantly associated with testing positive for SARS-CoV-2 were: contact with COVID-19 positive patient (OR 39.83 95%CI 17.52 to 90.55 p<0.0001), preexisting cardiac disease (OR 3.10 95%CI 1.19 to 5.02 p<0.0001), fever (OR 3.05 % 95% CI 1.67 to 5.58 p=0.0003), and anosmia/ageusia (OR 4.08 95%CI 1.69 to 9.84 p=0.002) (Table 3). Age between 2-9 years was negatively associated with testing positive for COVID-19, when taking the group of 10-18 years as reference (OR 0.33 CI 0.22 to 0.50 p<0.0001).

**Table 3.**
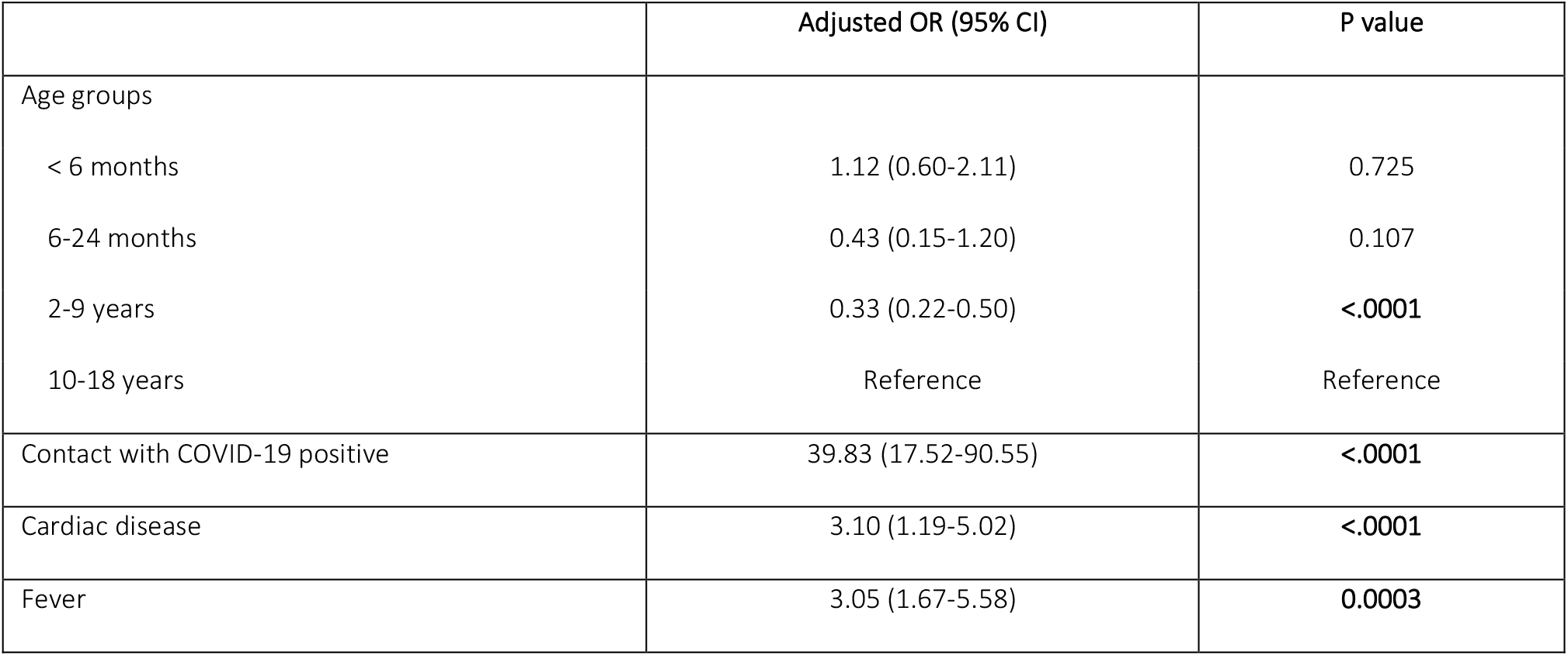
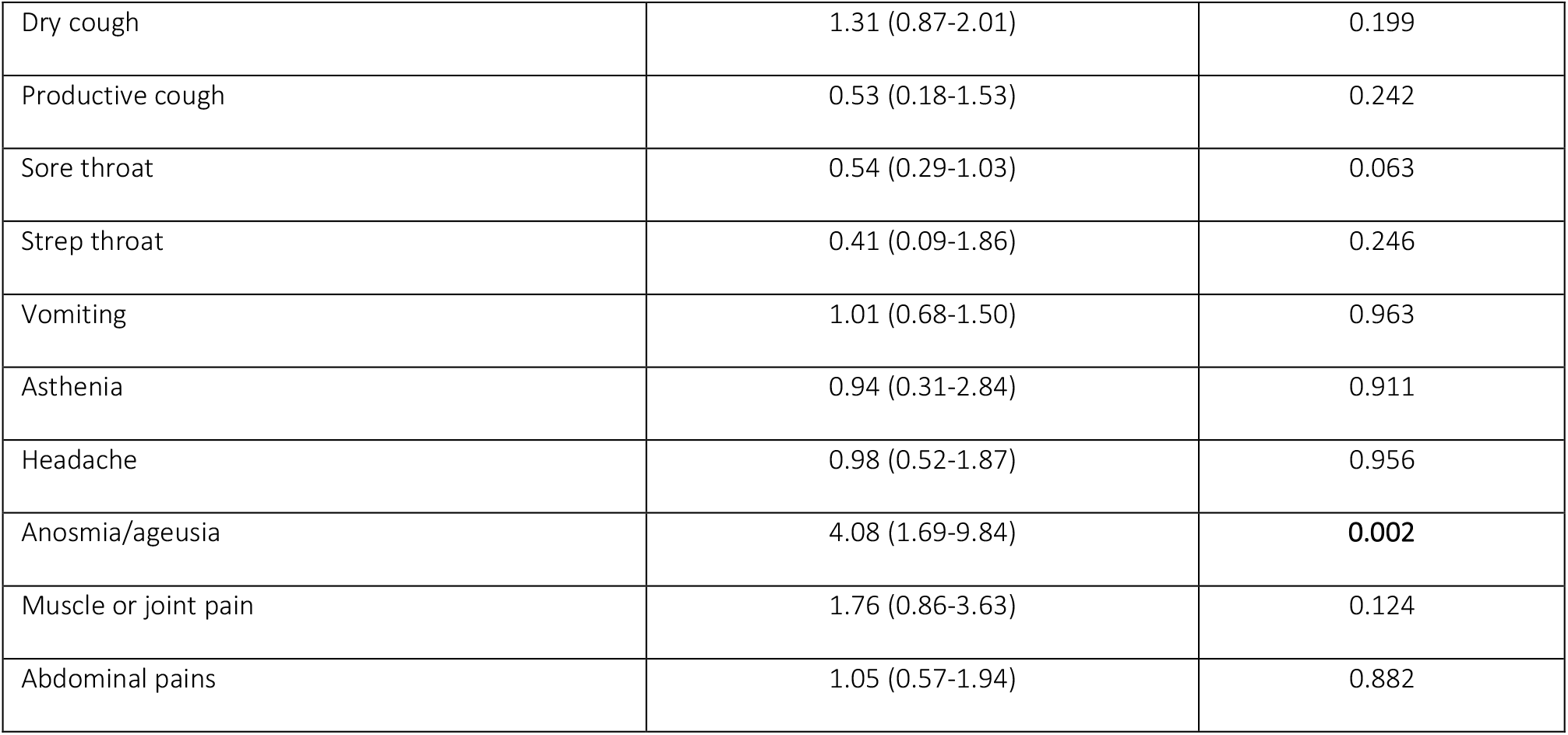
Multivariate analysis.

|  | Adjusted OR (95% CI) | P value |
| --- | --- | --- |
| Age groups |  |  |
| < 6 months | 1.12 (0.60-2.11) | 0.725 |
| 6-24 months | 0.43 (0.15-1.20) | 0.107 |
| 2-9 years | 0.33 (0.22-0.50) | **<.0001** |
| 10-18 years | Reference | Reference |
| Contact with COVID-19 positive | 39.83 (17.52-90.55) | **<.0001** |
| Cardiac disease | 3.10 (1.19-5.02) | **<.0001** |
| Fever | 3.05 (1.67-5.58) | **0.0003** |
| Dry cough | 1.31 (0.87-2.01) | 0.199 |
| Productive cough | 0.53 (0.18-1.53) | 0.242 |
| Sore throat | 0.54 (0.29-1.03) | 0.063 |
| Strep throat | 0.41 (0.09-1.86) | 0.246 |
| Vomiting | 1.01 (0.68-1.50) | 0.963 |
| Asthenia | 0.94 (0.31-2.84) | 0.911 |
| Headache | 0.98 (0.52-1.87) | 0.956 |
| Anosmia/ageusia | 4.08 (1.69-9.84) | **0.002** |
| Muscle or joint pain | 1.76 (0.86-3.63) | 0.124 |
| Abdominal pains | 1.05 (0.57-1.94) | 0.882 |

### Secondary analyses

Additional details on the 190 children testing positive for SARS-CoV-2 are reported in **Supplementary Table 3**. Overall, 139 (73.1%) children were cared for at home. The remaining were hospitalized in two types of wards: pediatric wards (55.6%) and general COVID-19 wards (53.3%). Cases treated with home care were either asymptomatic or had a mild or moderate presentation. Children received different types of treatments, with antibiotic, steroids and hydroxychloroquine being more frequently prescribed among hospitalized children (respectively 35.6% vs 10.5% p<0.001; 8.9% vs 0.0% p=0.006; 8.9% vs 0.0% p=0.006), while antipyretics/analgesic were much more frequently used in home care management (2.2% vs 51.8% p<0.001).

Socio-demographic, clinical characteristics and outcomes of children tested because of a COVID-19 positive contact and of those tested through hospital screening are reported in **Supplementary Table 4**. No significant difference was observed for any variable between the SARS-CoV-2 positives and the negatives in these two groups. None of the children positive for SARS-CoV-2 in these groups had respiratory distress, none required respiratory support, none were admitted in ICU, and all recovered.

No difference in disease severity were observed by age and sex, in children COVID −19 positive and symptoms suggestive of COVID-19 (**Supplementary Table 5**).

The rate of children testing positive for SARS-CoV-2 in each center was variable with an average rate of 9.17% (95%CI 0 to 18.71, **Supplementary Figure 2**). Specifically, in 17/20 (85%) centers the prevalence of COVID-19 positive swabs was below 15%, in two centers it was in between 15% to 20%, and in one center it was over 20%.

## DISCUSSION

This study adds to previous knowledge a description of characteristics and risk factors for SARS-CoV-2 among children. Notably, the clinical presentation of children with SARS-CoV-2 includes different possible scenarios. Besides the typical clinical picture with fever and respiratory signs or symptoms, this study suggested that COVID-19 in children may have gastrointestinal, neurological or cutaneous presentations, either in combination with other presentations or alone. These results are in line with reports from rheumatologists and dermatologists [25,26], gastroenterologists [27], neurologists and psychiatrists [28,29]. Additionally, emerging findings from COVID-19 screening among categories of people at risk, such as health workers, indicate that the current guidelines for testing may risk missing many cases [30]. Furthermore, it is important to acknowledge that, because the recommendations on SARS-Cov-2 case identification [24,31] indicate testing only for cases with either fever or respiratory signs, the real prevalence of other presentations (e.g., gastrointestinal, neurological and cutaneous) may have gone underestimated in this study, as well as in other studies. Guidelines for SARS-CoV-2 testing should be updated based on the evidence on clinical presentation of the disease in children and adults.

Our findings suggest that, in contrast with what has been observed in adults [18,19], in children there are very few features which help differentiate those affected by SARS-Cov-2 from those with other conditions. Specifically, some of the features identified so far in the few existing studies as predicting factors for COVID-19 in adults – such as obesity, leucopenia, lymphocytopenia, ground glass opacity at X-ray, and having both lungs affected [19], were not confirmed in children. This seems plausible, considering the generally mild presentation of SARS-CoV-2 in the pediatric age compared to adults, and the large number of other viruses which can affect children resulting in clinical pictures very similar to COVID-19.

Findings of this study indicate that a diagnosis of COVID-19 may be much more probable in those with contact with a person testing positive for SARS-Cov-2 (OR 39.83 95%CI 17.52 to 90.55), or with fever (OR 3.05 95%CI 1.67 to 5.58) or anosmia/ageusia (OR 4.08 95%CI 1.69 to 9.84 p=0.002). These results are in line with studies in adults [19, 28], and underscore the importance of testing all cases with exposure history and increased body temperature, as well as those with peculiar neurological signs.

Our findings related to young age as a protective factor (with children in the age range of 2-9 years being at lower risk of COVID-19 compared to reference group of 10-18 years, OR 0.33 95%CI 0.22 to 0.50) and to presence of cardiac disease as a risk factor (OR 3.10 95%CI 1.19 to 5.02) are novel, and warrant further confirmation and identification of causal mechanisms. Interestingly, the rate of comorbidities among all children accessing the health system with a presentation suggestive of COVID-19 was relatively high (355/2148, 16.5%). Nevertheless, the only comorbidity found to be associated with positive testing for SARS-CoV-2 was preexisting cardiac diseases. Interestingly, preexisting chronic kidney disease was found to be a significant predictive factor for COVID-19 diagnosis in one large study at the primary care level in England, not specific to children [18]. These results should be confirmed in larger studies in children. More studies should also contribute to explore if other factors apparently important in adults [18] – such as ethnicity, living situation, deprivation, children with smoking parents, obesity – may increase the risk of COVID-19 in children in Italy as well as in other settings.

This study suggests that COVID-19 is a mild disease in children in Italy: among the 190 children diagnosed with SARS-CoV-2 in our study, 12 (6.3%) had respiratory distress, only four (2.1%) required respiratory support, only two (1.1%) were admitted in ICU, and 100% recovered. These results are in line with surveillance data in Italy [32], and with previous reports on COVID-19 in children from different countries [4-6,16,33,34]. According to existing surveillance data from CDC, the number of deaths among children under 15 years of age with COVID-19 in United States was much lower than what was reported for children with seasonal influenzas in 2019-2020 (17 reported deaths for COVID-19 compared to 182 influenza-associated pediatric deaths) [35,36]. In contrast, data from adults indicate that COVID-19 may be more severe than influenzas in this population [37].

The sample of children hospitalized in this study is not negligible when compared with national data: at the end of the study period, Italian surveillance systems reported 227,204 confirmed SARS-CoV-2 cases in Italy, but only 123 hospitalized cases among children (i.e. age below 18 years) [32]. Our sample of 51 children hospitalized with COVID-19, thus, accounts for 41.4% of the total pediatric cases reported by national surveillance [32]. Clearly, larger prevalence studies as well as prospective longitudinal studies are needed to better understand the risk associated with COVID-19 in selected sub-populations of children at risk. Despite current preliminary evidence suggesting that even in children with underlying conditions – such as children with inflammatory bowel diseases [38], cancer [39], children in dialysis [40], and children with renal diseases in steroid treatment [41] - the risk of severe COVID-19 disease may be limited, much more solid data are needed.

This study highlights several interesting epidemiological findings, reporting the number of children tested in each center in the early phase of the pandemic and the rate of positivity. High heterogeneity across centers in the rate of positive SARS-CoV-2 testing is not surprising and may have multiple explanations. First, the epidemiology of the disease differs across Italy, where Regions in the North overall had a higher burden of cases compared to those in the South [42]. Second, case identification may have been affected by local protocols, testing capacities, and different implementation of testing recommendations, both at study start and over time. The number of total swabs per population has been reported as highly variable across Regions in Italy, and not always directly proportional to the incidence of COVID-19 disease, with considerable variations over time [43]. The implementation of case finding and contact tracing has been described as highly heterogenous in other countries [44] and would warrant further investigation to better interpret epidemiological curves. Epidemiological data on COVID-19, due also to limitations in the currently available technology for COVID-19 diagnosis (i.e., high rates of false negatives with nasal or nasopharyngeal swabs [45]) may not reflect the real incidence of the disease in each setting, and should, in general, be interpreted with extreme caution. Further studies should document knowledge, attitudes, and practices of case finding and contact tracing in Italy as well as in other settings. More accurate, acceptable, and sustainable tools are also needed for COVID-19 diagnosis.

Limitations of this study include the retrospective nature of data, possible selection bias toward more symptomatic cases due to the nature of the network, and the limitation in the technology currently available for COVID-19 diagnosis. Although the use of swabs is currently recommended as the “gold standard” for COVID-19 diagnosis, it has as major limitation of a high percentage of false negative cases [45]. Future studies, when better diagnostic tools will be available, should aim to confirm the observations of the present study. Strengths of this study include its pragmatic and descriptive nature, and the involvement of many pediatric centers in the national territory. More clinical and epidemiological studies are needed to further document the real incidence, presentation, risk factors and outcomes of children with COVID-19 infection in different pediatric subpopulations, to better characterize children at higher risk of the most severe forms of the disease.

## Data Availability

The authors confirm that the data supporting the findings of this study are available within the article and its supplementary materials.

## Data Sharing

De-identified individual participant data will be made available following completion of a data use agreement.

