## Supplementary material for "Characteristics and risk factors for SARS-CoV-2 among children in Italy: a cross-sectional study in 20 pediatric centers"

**Supplementary Table 1. Criteria for disease severity**

Asymptomatic: all the following must be present

1. No signs or symptoms
2. AND Negative Chest X-Ray
3. AND absence of criteria for other cases

Mild: any of the following (AND absence of criteria for more severe cases)

1. Symptoms of upper respiratory tract infection
2. AND absence of pneumonia at chest X-ray

Moderate: all the following (AND absence of criteria for more severe cases)

1. Cough or other symptom/sign AND pneumonia at chest X-ray
2. CT with lung lesions

Severe: any of the following (AND absence of criteria as for critical case)

1. Oxygen saturation <92% OR
2. Difficult breathing or other signs of severe respiratory distress (apnea, gasping, head nodding) or any other danger sign such as convulsion, restlessness, lethargy, severe dehydration, reduced reactivity, hypotonia
3. OR need for any respiratory support

Critical: Any of the following

1. Patient in ICU
2. OR mechanical ventilation
3. OR multiorgan failure
4. OR shock, encephalopathy, myocardial injury or heart failure, coagulation disfunction, acute kidney injury, coma.

Abbreviations: CT= Computed tomography; ICU = Intensive Care Unit;

Reference: Parri N, Magistà AM, Marchetti F, et al. Characteristic of COVID-19 infection in pediatric patients: early findings from two Italian Pediatric Research Networks [published online ahead of print, 2020 Jun 3]. Eur J Pediatr. 2020;1-9. doi:10.1007/s00431-020-03683-8 (adapted)

**Supplementary Table 2. Definition of tachypnea and tachycardia**

| **Tachypnea** | |
|---|---|
| Age | Respiratory rate (breaths/min) |
| 2 months | 60 |
| 2-12 months | 50 |
| 1-5 years | 40 |
| >5 years | 30 |
| **Tachycardia** | |
| Age | Heart rate (beats per min) |
| Term | >170 |
| 3-12 months | >165 |
| 1-5 years | >150 |
| 6-11 years | >135 |
| >11 years | >120 |

**Supplementary Table 3. Treatments and outcomes of children SARS-CoV-2 positive**

| | Groups of children | | | | | |
|---|---|---|---|---|---|---|
| | Symptoms suggestive for COVID-19 | | Contact with COVID-19 | | Screening for hospitalisation | |
| | Hospitalised N=45 n (%) | Home care N=114 n (%) | Hospitalised N=3 n (%) | Home care N=24 n (%) | Hospitalised N=3 n (%) | Home care N=1 ** n (%) |
| **Type of hospitalization*** | | | | | | |
| Pediatric ward | 25 (55.6) | - | 2 (66.7) | - | 1 | - |
| COVID-19 ward | 24 (53.3) | - | 2 (66.7) | - | 0 | - |
| In observation | 6 (13.3) | - | 0 | - | 1 (33.3) | - |
| Infectious diseases ward | 5 (11.1) | - | 0 | - | 0 | - |
| Intensive care unit | 2 (4.4) | - | 0 | - | 0 | - |
| **Type of respiratory support any** | 4 (2.5) | - | 0 | - | 0 | - |
| Oxygen | 3 (6.7) | - | 0 | - | 0 | - |
| High flow oxygen | 2 (4.4) | - | 0 | - | 0 | - |
| Non-invasive ventilation | 1 (2.2) | - | 0 | - | 0 | - |
| Mechanical ventilation | 0 | - | 0 | - | 0 | - |
| **Clinical course** | | | | | | |
| Asymptomatic | 6 (13.3)[1] | 2 (1.8) | 3 (100) | 24 (100) | 2 (66.7) | 1 (100) |
| Mild | 21 (46.7)[1] | 103 (90.4) | 0 | 0 | 0 | 0 |
| Moderate | 11 (24.4)[1] | 9 (7.9) | 0 | 0 | 0 | 0 |
| Severe | 5 (11.1)[1] | 0 | 0 | 0 | 1 (33.3) | 0 |
| Critical | 2 (4.4) | 0 | 0 | 0 | 0 | 0 |
| **Treatment** | | | | | | |
| Antibiotic | 16 (35.6)[1] | 12 (10.5) | 0 | 0 | 0 | 0 |
| Hydroxychloroquine | 4 (8.9)[1] | 0 | 0 | 0 | 0 | 0 |
| Steroids | 4 (8.9)[1] | 0 | 0 | 0 | 0 | 0 |
| Anticoagulant | 2 (4.4) | 0 | 0 | 0 | 0 | 0 |
| Antiviral | 2 (4.4) | 0 | 0 | 0 | 0 | 0 |
| Adrenaline | 2 (4.4) | 0 | 0 | 0 | 0 | 0 |
| Immunoglobulin | 2 (4.4) | 0 | 0 | 0 | 0 | 0 |
| Antipyretics/analgesic | 1 (2.2)[1] | 59 (51.8) | 0 | 0 | 0 | 0 |
| Others drugs | 1 (2.2) | 12 (10.5) | 0 | 1 (4.2) | 0 | 0 |
| Antiepileptic | 0 | 1 (0.9) | 0 | 0 | 0 | 0 |
| **Final outcome** | | | | | | |
| Cured | 45 (100) | 114 (100) | 3 (100) | 24 (100) | 3 (100) | 1 (100) |
| Referred | 0 | 0 | 0 | 0 | 0 | 0 |
| Dead | 0 | 0 | 0 | 0 | 0 | 0 |

[1] Significant difference ($p<0.05$) between hospitalised and home groups
* Total percentage exceeds 100% because some patients are double counted since moved from a ward to another.
** Child screened because of a day hospital
Abbreviations and symbols: IQR = inter-quartile ranges; - = not applicable.

**Supplementary Table 4. Characteristics of children tested for SARS-COv-2 because of contact with a positive case or within hospitals screening**

| | Contact with COVID-19 | | Screening for hospitalisation | |
|---|---|---|---|---|
| | **Positive swab N=27 n (%)** | **Negative swab N=25 n (%)** | **Positive swab N=4 n (%)** | **Negative swab N=290 n (%)** |
| **Age range** | 0-17 | 0-17 | 0-12 | 0-18 |
| **Age groups** | | | | |
| < 6 months | 2 (7.4) | 3 (12.0) | 1 (25.0) | 28 (9.7) |
| 6-24 months | 2 (7.4) | 1 (4.0) | 1 (25.0) | 36 (12.4) |
| 2-9 years | 10 (37.0) | 11 (44.0) | 1 (25.0) | 123 (42.4) |
| 10-18 years | 13 (48.1) | 10 (40.0) | 1 (25.0) | 99 (34.1) |
| Missing | 0 | 0 | 0 | 4 (1.4) |
| **Sex** | | | | |
| Male | 11 (40.7) | 11 (44.0) | 1 (25.0) | 186 (64.1) |
| Female | 16 (59.3) | 14 (56.0) | 3 (75.0) | 100 (34.5) |
| Missing | 0 | 0 | 0 | 4 (1.4) |
| **Contact with COVID-19 positive** | 27 (100) | 25 (100) | 0 | 3 (3.0) |
| **Relatives with respiratory symptoms** | 18 (66.7) | 19 (76.0) | 0 | 5 (1.7) |
| **Any co-morbidity** | 4 (14.8) | 1 (4.0) | 0 | 49 (16.9) |
| **Type of comorbidities** | | | | |
| Malformation, disability, neuromuscular diseases | 0 | 0 | 0 | 13 (4.5) |
| Cardiac disease | 0 | 0 | 0 | 5 (1.7) |
| Respiratory disease | 0 | 0 | 0 | 1 (0.3) |
| Asthma | 1 (3.7) | 0 | 0 | 0 |
| Primary immunodeficiency | 0 | 0 | 0 | 1 (0.3) |
| Secondary immunodeficiency | 0 | 0 | 0 | 1 (0.3) |
| Obesity | 2 (7.4) | 0 | 0 | 0 |
| Diabetes | 0 | 0 | 0 | 3 (1.0) |
| Psychiatric disorder | 0 | 0 | 0 | 9 (3.1) |
| Other | 1 (3.7) | 1 (4.0) | 0 | 19 (6.6) |
| **Disease severity at presentation** | | | | |
| Asymptomatic | 27 (100) | 25 (100) | 3 (75.0) | 135 (46.6) |
| Mild | 0 | 0 | 0 | 83 (28.6) |
| Moderate | 0 | 0 | 0 | 44 (15.2) |
| Severe | 0 | 0 | 1 (25.0) | 25 (8.6) |
| Critical | 0 | 0 | 0 | 3 (1.0) |
| **Symptoms and signs at presentation** | | | | |
| Fever | 0 | 0 | 0 | 22 (7.6) |
| **Respiratory symptoms, any** | 0 | 0 | 0 | 14 (4.8) |
| Respiratory distress | 0 | 0 | 0 | 5 (1.7) |
| Rhinorrhea | 0 | 0 | 0 | 4 (1.4) |
| Dry cough | 0 | 0 | 0 | 2 (0.7) |
| Productive cough | 0 | 0 | 0 | 0 |
| Sore throat | 0 | 0 | 0 | 2 (0.7) |
| Strep throat | 0 | 0 | 0 | 0 |
| Conjunctivitis | 0 | 0 | 0 | 2 (0.7) |
| Apnea | 0 | 0 | 0 | 0 |
| Thoracic pain | 0 | 0 | 0 | 2 (0.7) |
| **Gastrointestinal symptoms, any** | 0 | 0 | 0 | 19 (6.6) |
| Vomiting | 0 | 0 | 0 | 15 (5.2) |
| Diarrhea | 0 | 0 | 0 | 8 (2.8) |
| **Neurological symptoms, any** | 0 | 0 | 1 (25.0) | 18 (6.2) |
| Asthenia | 0 | 0 | 0 | 2 (0.7) |

| | | | | |
|---|---|---|---|---|
| Headache | 0 | 0 | 0 | 7 (2.4) |
| Anosmia/ageusia | 0 | 0 | 0 | 0 |
| Convulsion | 0 | 0 | 1 (25.0) | 7 (2.4) |
| Hyperactivity | 0 | 0 | 0 | 2 (0.7) |
| **Cutaneous symptoms, any** | 0 | 0 | 0 | 2 (0.7) |
| Skin manifestations | 0 | 0 | 0 | 2 (0.7) |
| Vasculitis | 0 | 0 | 0 | 0 |
| **Unspecific flu-like symptoms, any** | 0 | 0 | 0 | 18 (6.2) |
| Muscle or joint pain | 0 | 0 | 0 | 8 (2.8) |
| Nausea | 0 | 0 | 0 | 3 (1.0) |
| Inappetence | 0 | 0 | 0 | 10 (3.4) |
| Lymphadenitis | 0 | 0 | 0 | 2 (0.7) |
| **Other symptoms, any** | 0 | 0 | 0 | 61 (21.0) |
| Abdominal pains | 0 | 0 | 0 | 40 (13.8) |
| Oral manifestations (gingivostomatitis, aphthae) | 0 | 0 | 0 | 0 |
| Dental problems | 0 | 0 | 0 | 1 (0.3) |
| Urogenital disorders | 0 | 0 | 0 | 10 (3.4) |
| Ear problems | 0 | 0 | 0 | 1 (0.3) |
| Others | 0 | 0 | 0 | 11 (3.8) |
| **Vital parameters at presentation** | | | | |
| Tachycardia | 0/5 | 0/5 | 0/1 | 15/142 (10.6) |
| Tachypnea | 0/4 | 0/3 | 0/1 | 2/56 (3.6) |
| **Oxygen saturation level at presentation** | | | | |
| 91-92 | 0/5 | 0/7 | 0/1 | 0/121 |
| ≤90 | 0/5 | 0/7 | 0/1 | 0/121 |
| **Clinical examination at presentation** | | | | |
| **Lungs auscultations** | | | | |
| Negative | 7/9 (77.8) | 13/13 (100) | 1/1 (100) | 211/216 (97.7) |
| Crackles | 0/9 | 0/13 | 0/1 | 2/216 (0.9) |
| Wheezing | 0/9 | 0/13 | 0/1 | 0/216 |
| Absent breath sounds | 0/9 | 0/13 | 0/1 | 1/216 (0.5) |
| **Laboratory test**[1] | | | | |
| White blood cell count <5.5 (×109/L) | 0/2 | 0/1 | 0/1 | 2/62 (96.8) |
| Lymphocyte count <1.2 (×109/L) | 0/1 | 0/1 | 0 | 3/36 (8.3) |
| Neutrophil <1.50 (×109/L) | 0/2 | 0/1 | 0/1 | 3/57 (94.7) |
| C-reactive protein > 1 gr/dl | 0/2 | 0/1 | 0/1 | 22/52 (42.3) |
| Erythrocyte sedimentation rate > 20 mm/h | 0 | 0 | 0 | 3/5 (60.0) |
| Aspartate aminotransferase > 50 (U/L) | 0/1 | 0/1 | 0 | 3/23 (13.0) |
| Alanine aminotransferase > 45 (U/L) | 2/2 (100) | 0/1 | 0/1 | 4/49 (8.2) |
| D dimer >0.5 (μg/mL) | 0 | 0 | 0 | 1/1 (100) |
| **Chest X-ray** | 0 | 1 (4.0) | 0 | 8 (2.8) |
| Ground glass opacities [1] | 0 | 0 | 0 | 3/8 (37.5) |
| Negative [1] | 0 | 1 (4.0) | 0 | 4/8 (50.0) |
| Focal consolidation [1] | 0 | 0 | 0 | 1/8 (12.5) |
| Other description [1] | 0 | 0 | 0 | 0/8 |
| **Lung ultrasound** | 1 (3.7) | 0 | 0 | 0 |
| Ground glass opacities [1] | 0 | 0 | 0 | 0 |
| Negative [1] | 1 (3.7) | 0 | 0 | 0 |
| Focal consolidation [1] | 0 | 0 | 0 | 0 |
| Other description [1] | 0 | 0 | 0 | 0 |
| **CT scan** | 0 | 0 | 1 (25.0) | 0 |
| Ground glass opacities [1] | 0 | 0 | 1/1 (100) | 0 |

| | | | | |
|---|---|---|---|---|
| **Hospitalised[2]** | 3 (11.1) | 0 | 3 (75.0) | 272 (93.8) |
| **Respiratory support[1]** | 0 | 0 | 0 | 2 (0.7) |
| Oxygen | 0/3 | 0/0 | 0/3 | 0/272 |
| High flow oxygen | 0/3 | 0/0 | 0/3 | 0/272 |
| Noninvasive ventilation | 0/3 | 0/0 | 0/3 | 1/272 (0.4) |
| Mechanical ventilation | 0/3 | 0/0 | 0/3 | 2/272 (0.7) |
| **Cases in ICU** | 0/3 | 0/0 | 0/3 | 2/272 (0.7) |
| **Outcome** | | | | |
| Cured<br>Referred<br>Dead | 27 (100)<br>0<br>0 | 25 (100)<br>0<br>0 | 4 (100)<br>0<br>0 | 289 (99.7)<br>1(0.3)<br>0 |

No significant difference ($p<0.05$) between positive and negative swab in each group.

[1] Note: available in a subsample of cases

[2] Note: one child with social problem, one child with burns, one newborn

**Supplementary Table 5. Disease severity by sex and age in children COVID-19 positive and symptoms suggestive of COVID-19**

| **Disease severity** | **Sex** | |
|---|---|---|
| | **Male<br>N=77<br>n (%)** | **Female<br>N=82<br>n (%)** |
| Asymptomatic | 4 (5.2) | 4 (4.9) |
| Mild | 56 (72.7) | 68 (82.9) |
| Moderate | 13 (16.9) | 7 (8.5) |
| Severe | 3 (3.9) | 2 (2.4) |
| Critical | 1 (1.3) | 1 (1.2) |

| | **Age** | | | |
|---|---|---|---|---|
| | **< 6 months<br>N=19<br>n (%)** | **6-24 months<br>N=17<br>n (%)** | **2-9 years<br>N=37<br>n (%)** | **10-18 years<br>N=86<br>n (%)** |
| Asymptomatic | 3 (15.8) | 1 (5.9) | 2 (5.4) | 2 (2.3) |
| Mild | 11 (57.9) | 14 (82.4) | 32 (86.5) | 67 (77.9) |
| Moderate | 3 (15.8) | 1 (5.9) | 1 (2.7) | 15 (17.4) |
| Severe | 1 (5.3) | 1 (5.9) | 1 (2.7) | 2 (2.3) |
| Critical | 1 (5.3) | 0 | 1 (2.7) | 0 |

None significant difference ($p<0.05$) by age and sex

**Supplementary Figure 1. Geographical distribution of total cases (N=2494) and positive cases (N=190)**

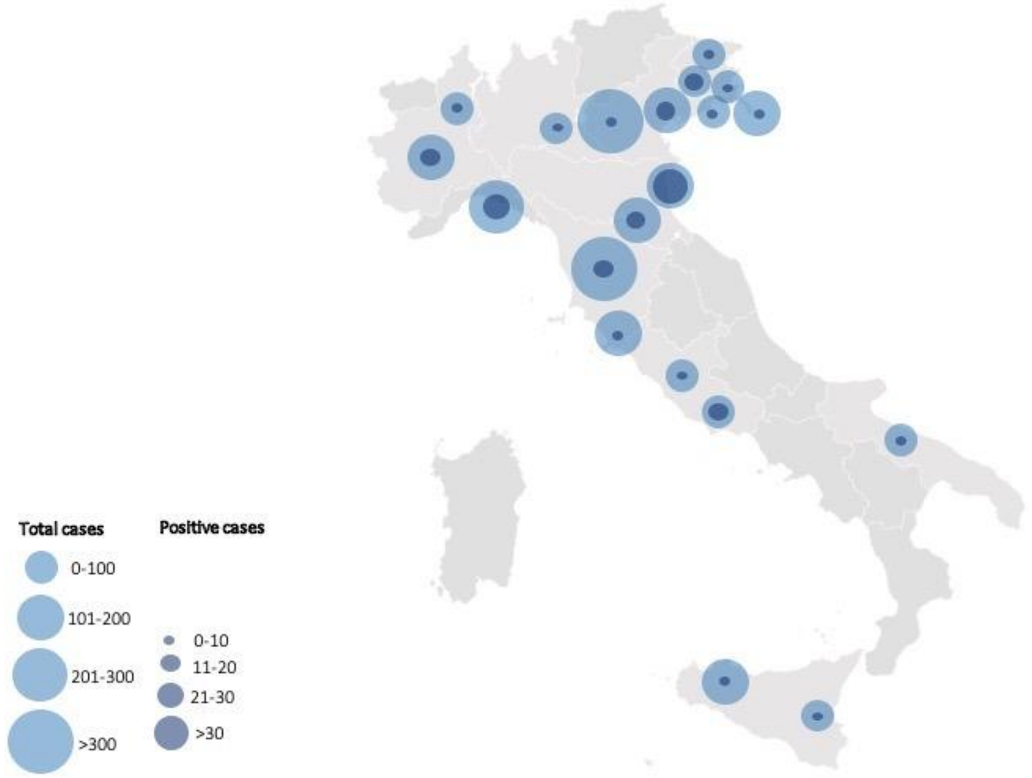

**Supplementary Figure 2. Percentage of SARS-CoV-2 positive cases across centers**

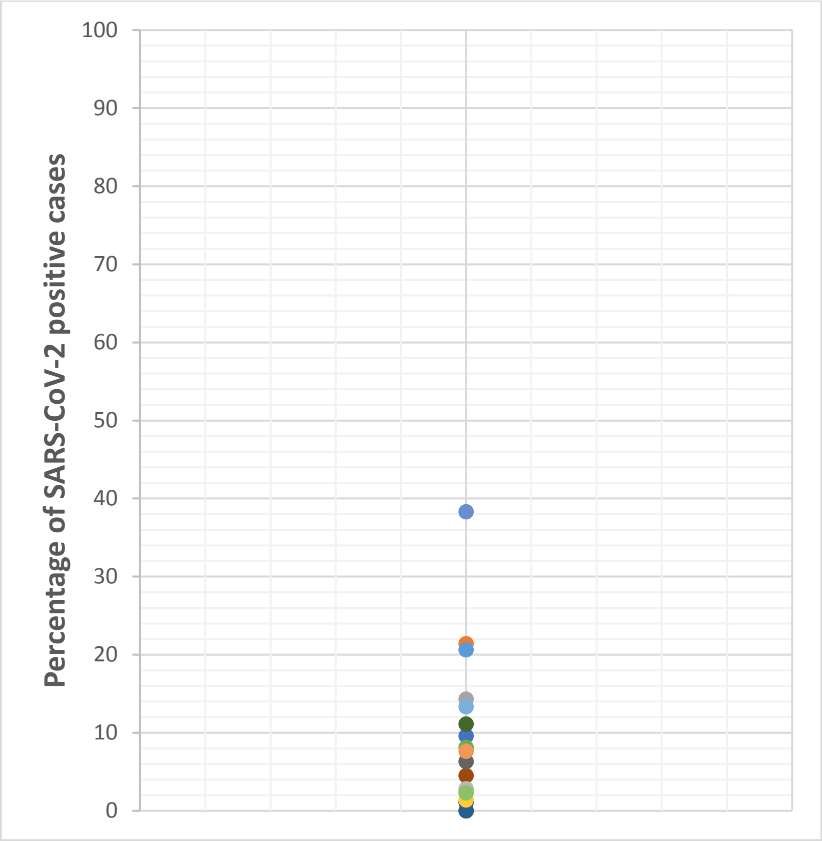
